# Cardiovascular Mortality in Patients with Gynecological Cancers: A Population-based Cohort Study

**DOI:** 10.1101/2024.10.13.24315340

**Authors:** Yue Yang, Jun-Ping Yang, Bing-Shu Li, Li-Wei Cheng, Shu-Jian Wei, Yu-Guo Chen

## Abstract

**Objective:** The survival rate of gynecological cancers (GCs) has improved significantly in recent decades. Patients with GCs did not necessarily succumb to the primary cancer. Cardiovascular health might be a critical determinant of long-term survival. This study aimed to investigate the mortality rate and risk factors associated with cardiovascular disease (CVD) death in patients with GCs.

**Methods:** A total of 399,399 cases of GCs diagnosed between 2000 and 2020 from the Surveillance, Epidemiology, and End Results (SEER) database were included in this study. The standardized mortality ratio (SMR) for CVD mortality was estimated. Prognostic factors for CVD death were assessed using cause-specific hazard ratios with 95% confidence intervals within a competing risk model, considering non-cardiovascular death as a competing risk.

**Results:** Of the 399,399 patients with GCs, 117,551 (29%) died from GCs, and 16,371 (4.1%) died from CVD. Of the CVD deaths, 73.2% were attributed to heart disease. The SMR of CVD mortality was highest in survivors diagnosed before age 45 years, and the risk of CVD mortality remained elevated throughout the follow-up period compared to the general United States (US) population. In recent years, the SMRs for CVD mortality risk increased steadily in all subtypes of GCs, except for vulvar cancer. Older age, black race, localized stage, unmarried/single/divorced, vaginal and vulvar cancers, and radiation therapy were associated with a higher risk of CVD mortality. A nomogram was developed and validated using these variables to predict CVD death risk in patients with GCs.

**Conclusions:** The risk of CVD mortality in patients with GCs was increased and was significantly higher compared with the general US population. A nomogram was constructed and validated to forecast the risk of CVD mortality in individuals with GCs. More attention should be paid to cardiovascular health during diagnosis to improve survival rates.

**HIGHLIGHTS:**

⇒ 29% of patients with gynecological cancers died from cancer, while 4.1% died from CVD, with 73.2% of CVD deaths attributed to heart disease.
⇒ In recent years, SMRs for CVD mortality increased steadily across all GC subtypes, except vulvar cancer.
⇒ Higher CVD mortality was linked to older age, black race, localized disease stage, unmarried/single/divorced status, and prior radiation therapy, especially in vaginal and vulvar cancers.
⇒ We developed and validated a nomogram to predict CVD death risk in GC patients, aiding personalized patient management.

## INTRODUCTION

Gynecologic cancers (GCs), including cancers of the cervix, ovaries, fallopian tubes, and uterus, accounted for approximately 11% of all cancers in women^1^. The overall incidence rate of GCs was second to breast cancer among women, posing a serious threat to their physical and mental health^2^. In the United States (US), the estimated number of new cases in 2022 was 65,950 for endometrial cancer, 19,880 for ovarian cancer, and 14,100 for cervical cancer^3^. In 2022, there were an estimated 12,810 deaths due to ovarian cancer and 32,830 deaths due to GCs in the US^3^.

The survival for advanced and recurrent GCs remained poor but advances in early detection, surgery, chemotherapy, and targeted therapies improved clinical outcomes^4–8^. However, the long-term survival of cancer patients has drawn attention to the impact of cardiovascular disease (CVD) as a significant contributor to mortality^9–11^. Cancer survivors were particularly vulnerable to CVD mortality due to the cardiotoxic effects of cancer treatments and common risk factors such as obesity, smoking, and alcohol consumption^12–14^. Cancer survivors had twice the risk of dying from CVD within the first year following a cancer diagnosis compared to the general US population^11^. This increased risk persisted throughout the survival period, highlighting the need for continued cardiovascular care in cancer survivors^11, 15^. In particular, survivors diagnosed with GCs faced an elevated risk of CVD mortality compared to the general US population^9, 10, 16^. Patients with endometrial or ovarian cancer were at a higher risk of dying from CVD than from the cancer itself, especially as time since diagnosis increased ^9, 10, 17–20^. Factors such as age, hypertension, and pre-existing heart conditions were associated with increased CVD death risk in patients with GCs^11, 21–23^.

Although CVD death was widely discussed for various malignancies, including breast cancer and lung cancer, the CVD mortality risk associated with GCs was not clearly described. To address this gap, we used the SEER data to determine the prevalence, trend, and risk factors for CVD death in patients with GCs.

## METHODS

### Data

This cohort study utilized data from the Surveillance, Epidemiology, and End Results (SEER) Registry. The SEER, representing nearly 28% of the US population, was an authoritative source of high-quality cancer registries worldwide, relying on systematic, standardized, and regular data collection procedures to ensure quality and prevent surveillance bias^24^. Ethics committee approval was waived because de-identified data were used. This report adhered to the Strengthening the Reporting of Observational Studies in Epidemiology (STROBE) guidelines for observational studies.

### Study Population

The International Classification of Diseases for Oncology, Third Edition (ICD-O-3) histology code was used to assign causes of death and cancer types. We defined CVD mortality due to cardiac disease, cerebrovascular disease, hypertension without cardiac disease, atherosclerosis, aortic aneurysm and dissection, and other diseases of the arteries, arterioles, and capillaries^25^. Data on patients diagnosed with GCs such as ovarian cancer, uterine cancer, cervical cancer, vaginal cancer, vulvar cancer, and others (malignant neoplasm of other and unspecified female genital organs, or malignant neoplasm of placenta) between 2000 and 2020 were extracted from the SEER database^26^. The exclusion criteria were as follows: (1) GCs were not the first malignancy; (2) patients were diagnosed only by autopsy or death certificate; (3) pediatric patients (age <18 years) were excluded from this study because diagnostic and therapeutic interventions might differ compared to adults; (4) causes of death were unknown.

Clinical variables of interest included age at diagnosis, race, year of diagnosis, stage, grade, marital status, cancer type, and initial treatment regimen. Age at diagnosis was divided into three categories: less than 45, 45 to 65, and 65 years and older. Race was recorded as White, Black, and Other. The calendar years at diagnosis were categorized as 2000 to 2005, 2005 to 2010, 2010 to 2015, and 2015 to 2020. Tumor stage was classified as localized, regional, and distant. Tumor grade was divided into grade I (well differentiated), grade II (moderately differentiated), grade III (poorly differentiated), and grade IV (undifferentiated). Marital status was categorized as “married” and “unmarried/single/divorced”. Patients were divided into the following treatment categories: surgery and not/unknown, chemotherapy and not/unknown, and radiotherapy and not/unknown.

### Statistical Analysis

Categorical variables in baseline characteristics were assessed using the chi-square test. To further evaluate the interaction between GCs and CVD mortality, we calculated the standardized mortality ratios (SMRs) to quantify the risk of CVD death in GC patients. The SMR was calculated as the ratio of observed deaths to expected deaths^27^. Joinpoint regression was used to calculate average annual percentage changes (AAPC) for evaluating trends over time^28^.

CVD-specific mortality hazard ratios (HRs) with corresponding 95% confidence intervals (CIs) were calculated to estimate the relative association between individual risk and prognostic factors for CVD mortality using univariate and multivariate competing risk models (with non-cardiovascular death as a competing risk)^29, 30^. The classical proportional hazard model examined the effect of covariates on the cause-specific hazard function but ignored the competing nature of multiple causes for the same event, leading to inaccurate evaluation of variables in the marginal probability analysis for cause-specific events^30^. Fine and Gray developed a survival analysis method using the inverse probability censoring weighting technique with a time-dependent weight function to correctly estimate the marginal probability of an event in the presence of competing events^31, 32^. This competing risk model was used to determine whether baseline clinical characteristics and treatment strategies were associated with CVD death. Independent variables included age at diagnosis, race, tumor grade, stage, and initial treatments such as surgery, chemotherapy, and radiation^33^.

Seventy percent of patients with GCs were randomly assigned to the discovery cohort and the remaining 30 percent to the validation cohort. A nomogram was created based on the multivariate analysis results^34^. The performances of the nomogram in the discovery and validation cohorts were evaluated as follows: The area under the receiver operating characteristic curve (AUC) was calculated to evaluate the discriminative ability of the nomogram ^35^. An AUC value above 0.7 was considered indicative of good predictive ability^36^. The concordance index (C-index) was used to evaluate the predictive performance of the nomogram^37^. A calibration plot was used to compare the actual and predicted values and assess the model’s consistency ^38^. Better model accuracy was indicated when the calibration curve aligned with the 45° reference line ^39^. The model’s clinical validity was evaluated through decision-curve analysis (DCA), where the ordinate represented threshold probability and the abscissa corresponded to net benefit^40^. A higher DCA curve indicated a greater net benefit for the model^40^.

The analysis was conducted using SEER*Stat 8.3.6 (National Cancer Institute, USA), Microsoft Excel 15.0.4 (Microsoft, Redmond, WA), GraphPad Prism 5 (GraphPad Software Inc., San Diego, CA, USA), Joinpoint Regression Program 5.0.2 (Variance Covariance Matrix Beta), and R 3.4.4 software (Vienna, Austria). Two-tailed hypothesis tests were performed, with statistical significance at *P* < 0.05^41^.

## RESULTS

### Patient Characteristics

From 2000 to 2020, a total of 399,399 patients with GCs were enrolled in the SEER database, including 94,563 patients (24%) with ovarian cancer, 210,664 patients (53%) with uterine cancer, 64,025 patients (16%) with cervical cancer, 16,370 patients (4.1%) with vulvar cancer, and 4,176 patients (1.0%) with vaginal cancer. Most tumors occurred in white patients (80%). A total of 202,153 patients with GCs (51%) had localized cancer, and 98,995 patients (25%) had distant cancer. The patients’ information was presented in **Table 1**.

**Table 1.** Participant Characteristics. Abbreviations: CVD: cardiovascular disease. ^a^ malignant neoplasm of other and unspecified female genital organs, or malignant neoplasm of placenta.

| Characteristics | Gynecological cancer | Ovarian cancer | Uterine cancer | Cervical cancer | Vaginal cancer | Vulvar cancer | Other <sup>a</sup> |
| --- | --- | --- | --- | --- | --- | --- | --- |
| Age at diagnosis |  |  |  |  |  |  |  |
| <40 | 35,474 (8.9%) | 7,222 (7.6%) | 8,686 (4.1%) | 17,738 (28%) | 870 (5.3%) | 160 (3.8%) | 798 (8.3%) |
| 40-65 | 152,431 (38%) | 33,772 (36%) | 79,842 (38%) | 29,877 (47%) | 4,935 (30%) | 1,234 (30%) | 2,771 (29%) |
| ≥65 | 211,494 (53%) | 53,569 (57%) | 122,136 (58%) | 16,410 (26%) | 10,565 (65%) | 2,782 (67%) | 6,032 (63%) |
| Attained age |  |  |  |  |  |  |  |
| <40 | 15,093 (3.8%) | 3,829 (4.0%) | 3,158 (1.5%) | 7,286 (11%) | 301 (1.8%) | 66 (1.6%) | 453 (4.7%) |
| 40-65 | 99,161 (25%) | 23,758 (25%) | 39,024 (19%) | 30,477 (48%) | 3,048 (19%) | 808 (19%) | 2,046 (21%) |
| ≥65 | 285,145 (71%) | 66,976 (71%) | 168,482 (80%) | 26,262 (41%) | 13,021 (80%) | 3,302 (79%) | 7,102 (74%) |
| Race |  |  |  |  |  |  |  |
| White | 321,910 (81%) | 78,069 (83%) | 169,829 (81%) | 48,645 (76%) | 14,306 (87%) | 3,249 (78%) | 7,812 (81%) |
| Black | 39,965 (10%) | 7,677 (8.1%) | 20,905 (9.9%) | 8,555 (13%) | 1,345 (8.2%) | 621 (15%) | 862 (9.0%) |
| Other | 37,524 (9.4%) | 8,817 (9.3%) | 19,930 (9.5%) | 6,825 (11%) | 719 (4.4%) | 306 (7.3%) | 927 (9.7%) |
| Stage |  |  |  |  |  |  |  |
| Localized | 202,153 (51%) | 18,139 (19%) | 142,407 (68%) | 29,187 (46%) | 9,741 (60%) | 1,288 (31%) | 1,391 (14%) |
| Regional | 79,546 (20%) | 10,416 (11%) | 37,938 (18%) | 23,134 (36%) | 4,737 (29%) | 1,628 (39%) | 1,693 (18%) |
| Distant metastasis | 98,995 (25%) | 59,970 (63%) | 22,616 (11%) | 8,784 (14%) | 990 (6.0%) | 834 (20%) | 5,801 (60%) |
| None/unknown | 18,705 (4.7%) | 6,038 (6.4%) | 7,703 (3.7%) | 2,920 (4.6%) | 902 (5.5%) | 426 (10%) | 716 (7.5%) |
| Grade |  |  |  |  |  |  |  |
| Grade I | 72,291 (18%) | 5,496 (5.8%) | 58,271 (28%) | 5,355 (8.4%) | 2,764 (17%) | 256 (6.1%) | 149 (1.6%) |
| Grade II | 70,927 (18%) | 9,654 (10%) | 39,684 (19%) | 16,202 (25%) | 4,048 (25%) | 910 (22%) | 429 (4.5%) |
| Grade III | 72,608 (18%) | 24,047 (25%) | 28,034 (13%) | 15,631 (24%) | 1,760 (11%) | 1,059 (25%) | 2,077 (22%) |
| Grade IV | 25,162 (6.3%) | 11,411 (12%) | 10,542 (5.0%) | 1,341 (2.1%) | 119 (0.7%) | 133 (3.2%) | 1,616 (17%) |
| Unknown | 158,411 (40%) | 43,955 (46%) | 74,133 (35%) | 25,496 (40%) | 7,679 (47%) | 1,818 (44%) | 5,330 (56%) |
| Marital status |  |  |  |  |  |  |  |
| Married | 191,226 (48%) | 45,557 (48%) | 105,911 (50%) | 27,039 (42%) | 6,281 (38%) | 1,555 (37%) | 4,883 (51%) |
| Unmarried/Single/Divorced | 187,879 (47%) | 45,164 (48%) | 94,140 (45%) | 33,243 (52%) | 8,687 (53%) | 2,309 (55%) | 4,336 (45%) |
| None/unknown | 20,294 (5.1%) | 3,842 (4.1%) | 10,613 (5.0%) | 3,743 (5.8%) | 1,402 (8.6%) | 312 (7.5%) | 382 (4.0%) |
| Year at diagnosis |  |  |  |  |  |  |  |
| 2000-2004 | 83,169 (21%) | 22,777 (24%) | 38,720 (18%) | 16,010 (25%) | 3,520 (22%) | 996 (24%) | 1,146 (12%) |
| 2005-2009 | 88,677 (22%) | 22,995 (24%) | 44,377 (21%) | 15,316 (24%) | 3,609 (22%) | 953 (23%) | 1,427 (15%) |
| 2010-2014 | 98,200 (25%) | 22,898 (24%) | 53,410 (25%) | 14,707 (23%) | 3,980 (24%) | 1,027 (25%) | 2,178 (23%) |
| 2015-2020 | 129,353 (32%) | 25,893 (27%) | 74,157 (35%) | 17,992 (28%) | 5,261 (32%) | 1,200 (29%) | 4,850 (51%) |
| Surgery |  |  |  |  |  |  |  |
| None/unknown | 79,176 (20%) | 23,112 (24%) | 19,749 (9.4%) | 28,042 (44%) | 3,131 (19%) | 2,783 (67%) | 2,359 (25%) |
| Yes | 320,223 (80%) | 71,451 (76%) | 190,915 (91%) | 35,983 (56%) | 13,239 (81%) | 1,393 (33%) | 7,242 (75%) |
| Radiation |  |  |  |  |  |  |  |
| None/unknown | 301,928 (76%) | 93,153 (99%) | 156,001 (74%) | 29,899 (47%) | 12,213 (75%) | 1,398 (33%) | 9,264 (96%) |
| Yes | 97,471 (24%) | 1,410 (1.5%) | 54,663 (26%) | 34,126 (53%) | 4,157 (25%) | 2,778 (67%) | 337 (3.5%) |
| Chemotherapy |  |  |  |  |  |  |  |
| None/unknown | 257,678 (65%) | 33,607 (36%) | 170,596 (81%) | 34,566 (54%) | 13,833 (85%) | 2,272 (54%) | 2,804 (29%) |
| Yes | 141,721 (35%) | 60,956 (64%) | 40,068 (19%) | 29,459 (46%) | 2,537 (15%) | 1,904 (46%) | 6,797 (71%) |
| Treatment |  |  |  |  |  |  |  |
| Surgery only | 177,046 (44%) | 19,752 (21%) | 122,569 (58%) | 22,272 (35%) | 10,566 (65%) | 543 (13%) | 1,344 (14%) |
| Radiation only | 7,876 (2.0%) | 121 (0.1%) | 2,849 (1.4%) | 3,613 (5.6%) | 553 (3.4%) | 676 (16%) | 64 (0.7%) |
| Chemotherapy only | 14,378 (3.6%) | 9,172 (9.7%) | 2,655 (1.3%) | 1,478 (2.3%) | 102 (0.6%) | 116 (2.8%) | 855 (8.9%) |
| Chemotherapy and radiation | 21,889 (5.5%) | 233 (0.2%) | 1,370 (0.7%) | 17,794 (28%) | 1,071 (6.5%) | 1,320 (32%) | 101 (1.1%) |
| Surgery and radiation | 37,723 (9.4%) | 148 (0.2%) | 32,303 (15%) | 3,524 (5.5%) | 1,309 (8.0%) | 382 (9.1%) | 57 (0.6%) |
| Surgery and chemotherapy | 75,471 (19%) | 50,643 (54%) | 17,902 (8.5%) | 992 (1.5%) | 140 (0.9%) | 68 (1.6%) | 5,726 (60%) |
| Surgery, radiation and chemotherapy | 29,983 (7.5%) | 908 (1.0%) | 18,141 (8.6%) | 9,195 (14%) | 1,224 (7.5%) | 400 (9.6%) | 115 (1.2%) |
| None/unknown | 35,033 (8.8%) | 13,586 (14%) | 12,875 (6.1%) | 5,157 (8.1%) | 1,405 (8.6%) | 671 (16%) | 1,339 (14%) |
| Cause of death |  |  |  |  |  |  |  |
| Alive | 236,258 (59%) | 36,594 (39%) | 144,536 (69%) | 40,074 (63%) | 8,484 (52%) | 1,619 (39%) | 4,951 (52%) |
| CVD | 16,371 (4.1%) | 2,569 (2.7%) | 10,178 (4.8%) | 1,708 (2.7%) | 1,434 (8.8%) | 271 (6.5%) | 211 (2.2%) |
| Cancer-index | 117,551 (29%) | 50,821 (54%) | 38,432 (18%) | 18,362 (29%) | 4,096 (25%) | 1,848 (44%) | 3,992 (42%) |
| Cancer-non-index | 6,984 (1.7%) | 1,021 (1.1%) | 4,156 (2.0%) | 1,040 (1.6%) | 550 (3.4%) | 116 (2.8%) | 101 (1.1%) |
| None-CVD other cause | 22,235 (5.6%) | 3,558 (3.8%) | 13,362 (6.3%) | 2,841 (4.4%) | 1,806 (11%) | 322 (7.7%) | 346 (3.6%) |

Abbreviations: CVD: cardiovascular disease

### CVD Death

The cumulative mortality for causes of death in patients with GCs was shown in **Figure 1**. The highest cumulative mortality was due to cancer-specific deaths (72.1%), followed by non-CVD causes (13.6%) and CVD causes (10.0%). Vaginal cancer patients (18.2%) had the highest risk of dying from CVD, followed by patients with uterine cancer (15.4%) and vulvar cancer (10.6%) (**Figure 1**). Among non-cancer deaths in patients with GCs, CVD mortality was the most common cause (41.1%). The majority (73.2%) of CVD deaths were due to heart disease (**Figure 1**). We observed that increasing age at diagnosis was associated with an increased percentage of cancer patients dying from index cancer and cardiovascular disease. The CVD death rate in patients younger than age 40 years was 1.1% at 20 years, compared with 14.0% in patients aged 65 years or older (**Table S1**). We also observed a trend indicating that a greater percentage of cancer patients who were recently diagnosed had a higher chance of developing cardiovascular disease and experiencing its consequent effects.

**Figure 1.**
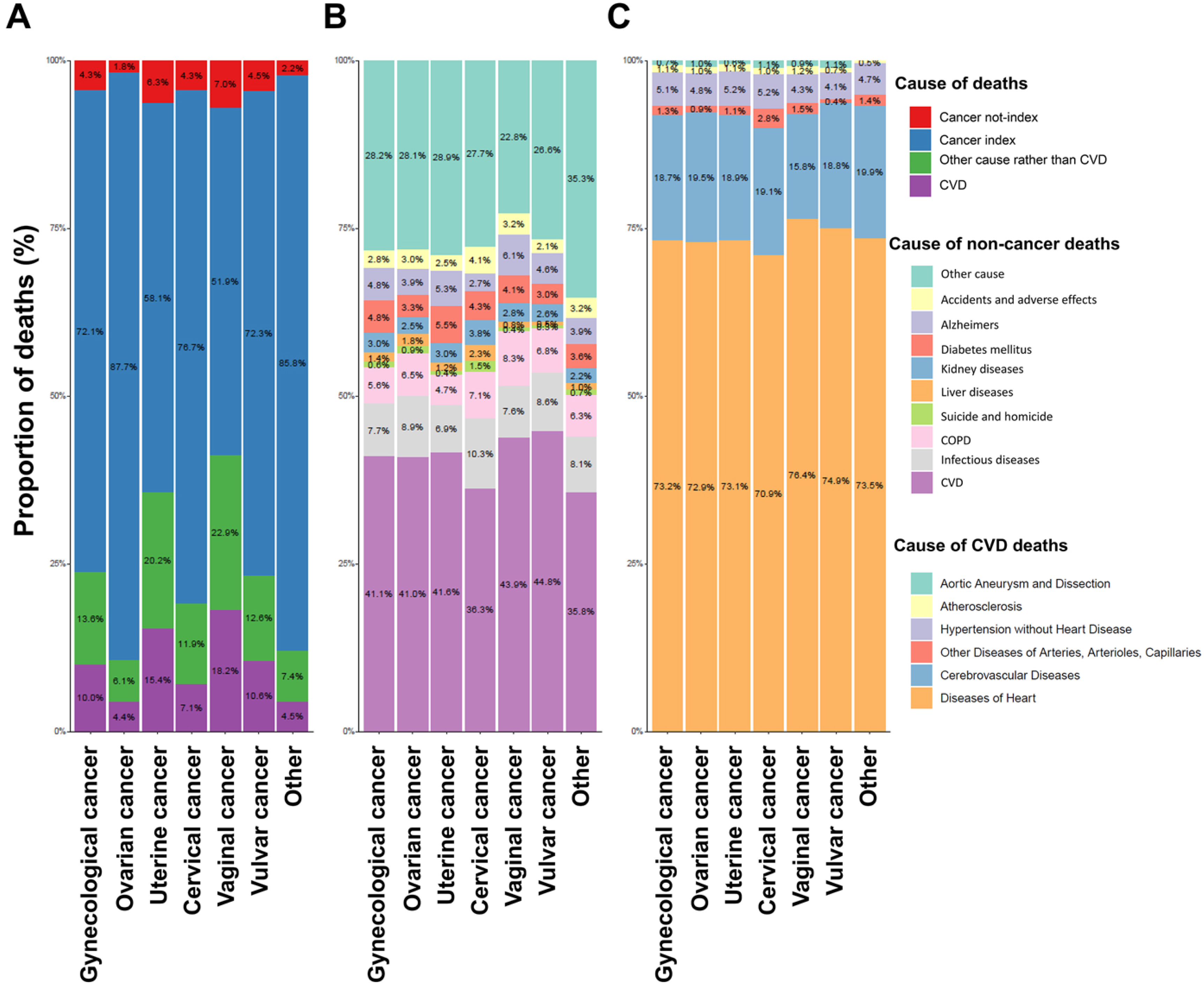
Distribution of the Most Common Causes of Death in Patients with Gynecological Cancers.

The SMR of CVD mortality was highest in survivors diagnosed under the age of 45 years compared to the general US population. The younger a cancer survivor was at diagnosis, the higher their risk of dying from cardiovascular disease. Survivors diagnosed with GCs before age 45 had a risk of CVD mortality more than 2.12 times higher than the general US population. The risk of CVD death among cancer survivors gradually decreased with increasing age at diagnosis (**Figure 2** and **Table 2**). The risk of CVD mortality remained elevated throughout the follow-up period compared to the general US population (**Figure 2**). In recent years, the SMR of CVD death increased in survivors with ovarian cancer (AAPC = 0.0668, 95% CI = 0.0537 to 0.0798), uterine cancer (AAPC = 0.0229, 95% CI = 0.0184 to 0.0274), cervical cancer (AAPC = 0.0417, 95% CI = 0.027 to 0.0564), and vaginal cancer (AAPC = 0.1071, 95% CI = 0.0662 to 0.1479), except vulvar cancer (AAPC = -0.0239, 95% CI = -0.0413 to -0.0064) (**Figure 2** and **Table S2**).

**Figure 2.**
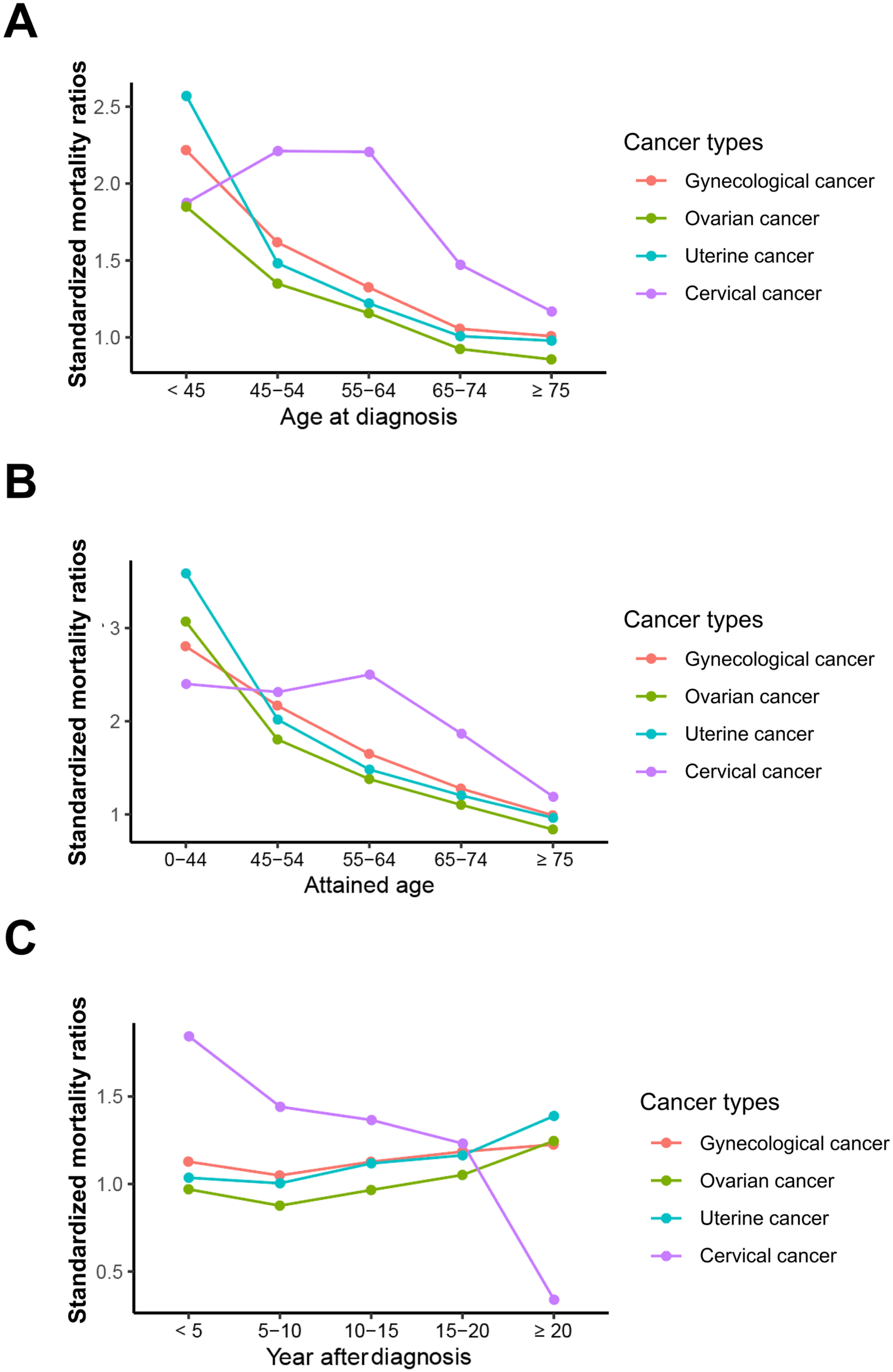
Standardized Mortality Ratios in Patients with Gynecological Cancers by (A) Age at Diagnosis, (B) Attained Age, and (C) Time After Diagnosis.

**Figure 3.**
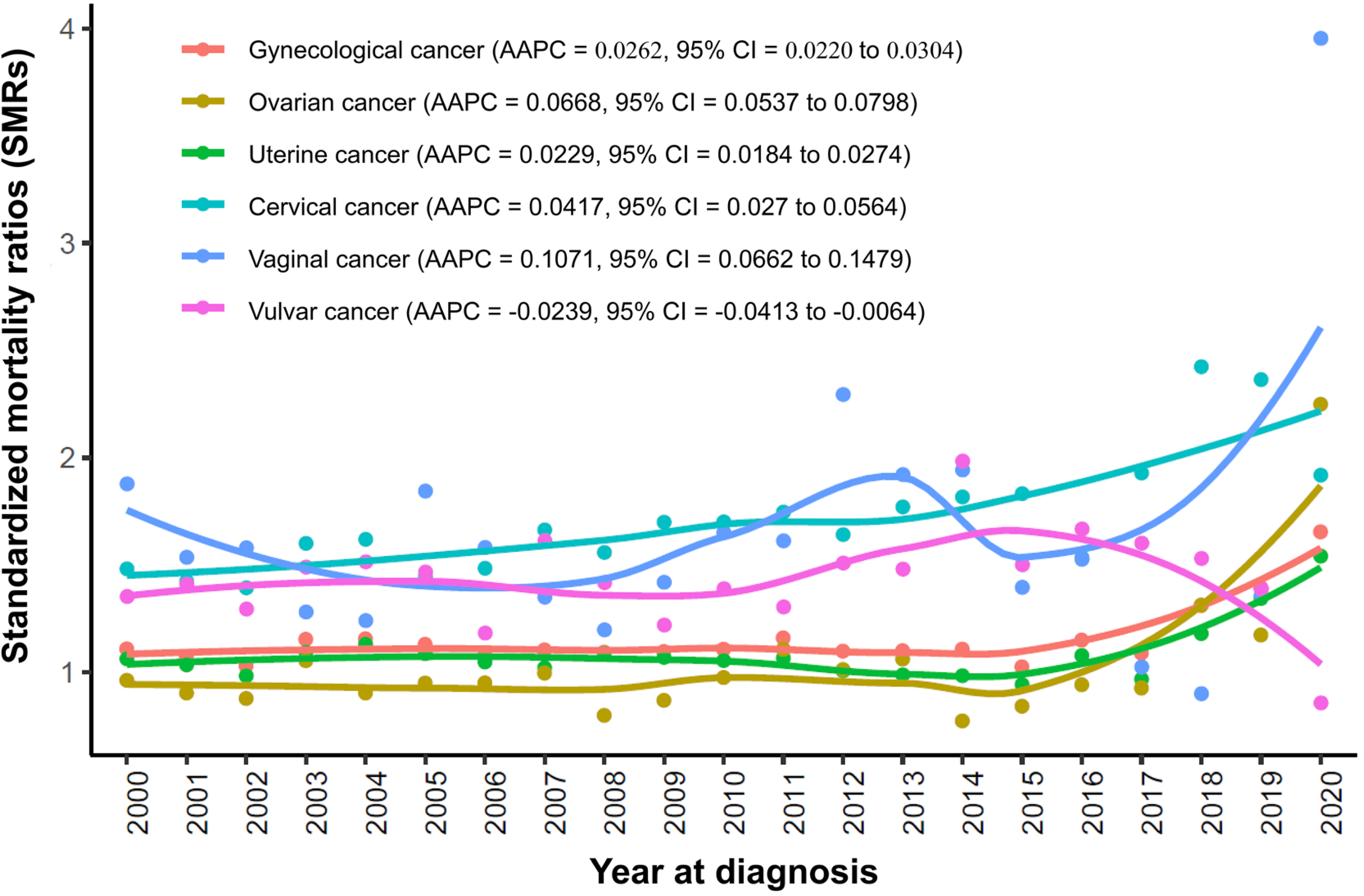
Trends in CVD Death by Calendar Years (2010–2020) among Survivors Diagnosed with Gynecological Cancers by Cancer Types. *Abbreviations: CVD: cardiovascular disease; AAPC: average annual percentage change;*

**Table 2.** Standardized Mortality Ratios in Patients with Gynecological Cancers. The SMRs were estimated as the ratios of observed to expected number of deaths. The observed values represented the number of CVD deaths in patients with gynecological cancers, and the expected values represented the number of individuals who died of CVD in the US general population, with a similar distribution of age, sex, race, and calendar year. *Abbreviations: SMR: standardized mortality ratio; CI: confidence interval; ^a^ * suggested that the 95% confidence interval did not cross 1.0 and the SMR was significant. ^b^ Reference population: the general US population based on the 2000 US standard population, Patients with multiple primary tumors were excluded automatically*.

| Characteristics | Observed | Expected | SMR (95% CI)<sup>a, b</sup> |
| --- | --- | --- | --- |
| **Age at diagnosis** |  |  |  |
| < 45 | 383 | 173 | 2.218 (2.001-2.451) * |
| 45-54 | 1024 | 633 | 1.618 (1.521-1.721) * |
| 55-64 | 2616 | 1974 | 1.325 (1.275-1.377) * |
| 65-74 | 4103 | 3886 | 1.056 (1.024-1.088) * |
| ≥ 75 | 7311 | 7253 | 1.008 (0.985-1.031) |
| **Race** |  |  |  |
| White | 12857 | 12131 | 1.06 (1.042-1.078) * |
| Black | 1722 | 1217 | 1.415 (1.348-1.483) * |
| Other | 858 | 571 | 1.502 (1.404-1.606) * |
| **Stage** |  |  |  |
| Localized | 5524 | 5591 | 0.988 (0.962-1.014) |
| Regional | 2500 | 1979 | 1.263 (1.214-1.314) * |
| Distant metastasis | 1166 | 977 | 1.194 (1.126-1.264) * |
| **Marital status** |  |  |  |
| Unmarried/Single/Divorced | 9132 | 7293 | 1.252 (1.227-1.278) * |
| Married | 5412 | 5950 | 0.910 (0.885-0.934) |
| **Grade** |  |  |  |
| Grade I | 3894 | 3870 | 1.006 (0.975-1.038) |
| Grade II | 4056 | 3655 | 1.110 (1.076-1.145) * |
| Grade III | 3030 | 2664 | 1.137 (1.097-1.179) * |
| Grade IV | 686 | 705 | 0.973 (0.902-1.049) * |
| **Year at diagnosis** |  |  |  |
| 2000-2004 | 6646 | 6036 | 1.101 (1.075-1.128) * |
| 2005-2009 | 4464 | 4052 | 1.102 (1.069-1.134) * |
| 2010-2014 | 2925 | 2620 | 1.116 (1.076-1.158) * |
| 2015-2020 | 1402 | 1211 | 1.158 (1.098-1.22) * |
| **Cancer type** |  |  |  |
| Gynecological cancer | 15437 | 13919 | 1.109 (1.092-1.127) * |
| Ovarian cancer | 2123 | 2233 | 0.951 (0.911-0.992) |
| Uterine cancer | 9711 | 9219 | 1.053 (1.032-1.074) * |
| Cervical cancer | 1632 | 1032 | 1.582 (1.506-1.660) * |
| Vaginal cancer | 257 | 165 | 1.560 (1.375-1.762) * |
| Vulvar cancer | 1387 | 968 | 1.432 (1.358-1.51) * |
| **Surgery** |  |  |  |
| None/unknown | 3329 | 1451 | 2.294 (2.217-2.374) * |
| Yes | 12108 | 12468 | 0.971 (0.954-0.989) |
| **Chemotherapy** |  |  |  |
| None/unknown | 12568 | 11069 | 1.135 (1.116-1.155) * |
| Yes | 2869 | 2850 | 1.007 (0.97-1.044) |
| **Radiation** |  |  |  |
| None/unknown | 11329 | 10521 | 1.077 (1.057-1.097) * |
| Yes | 4108 | 3398 | 1.209 (1.172-1.247) * |

### Factors Associated with CVD Mortality

We assessed factors associated with CVD mortality using a multivariable competing risk model (with non-cardiovascular death as a competing risk). In patients with GCs, the mortality was higher with increasing age (> 65 vs. < 40, hazard ratio [HR] 15.3, 95% CI 13.1–17.9; *P* < 0.001). Black women were more likely to die due to cardiovascular disease (HR 1.12, 95% CI 1.07-1.18; *P* < 0.001), as were unmarried/single/divorced individuals (HR 1.57, 95% CI 1.52-1.62; *P* < 0.001). Patients treated with surgery (HR 0.64, 95% CI 0.61-0.67; *P* < 0.001) or chemotherapy (HR 0.59, 95% CI 0.56-0.62; *P* < 0.001) had lower CVD mortality. (**Figure 4** and **Table 3**).

**Figure 4.**
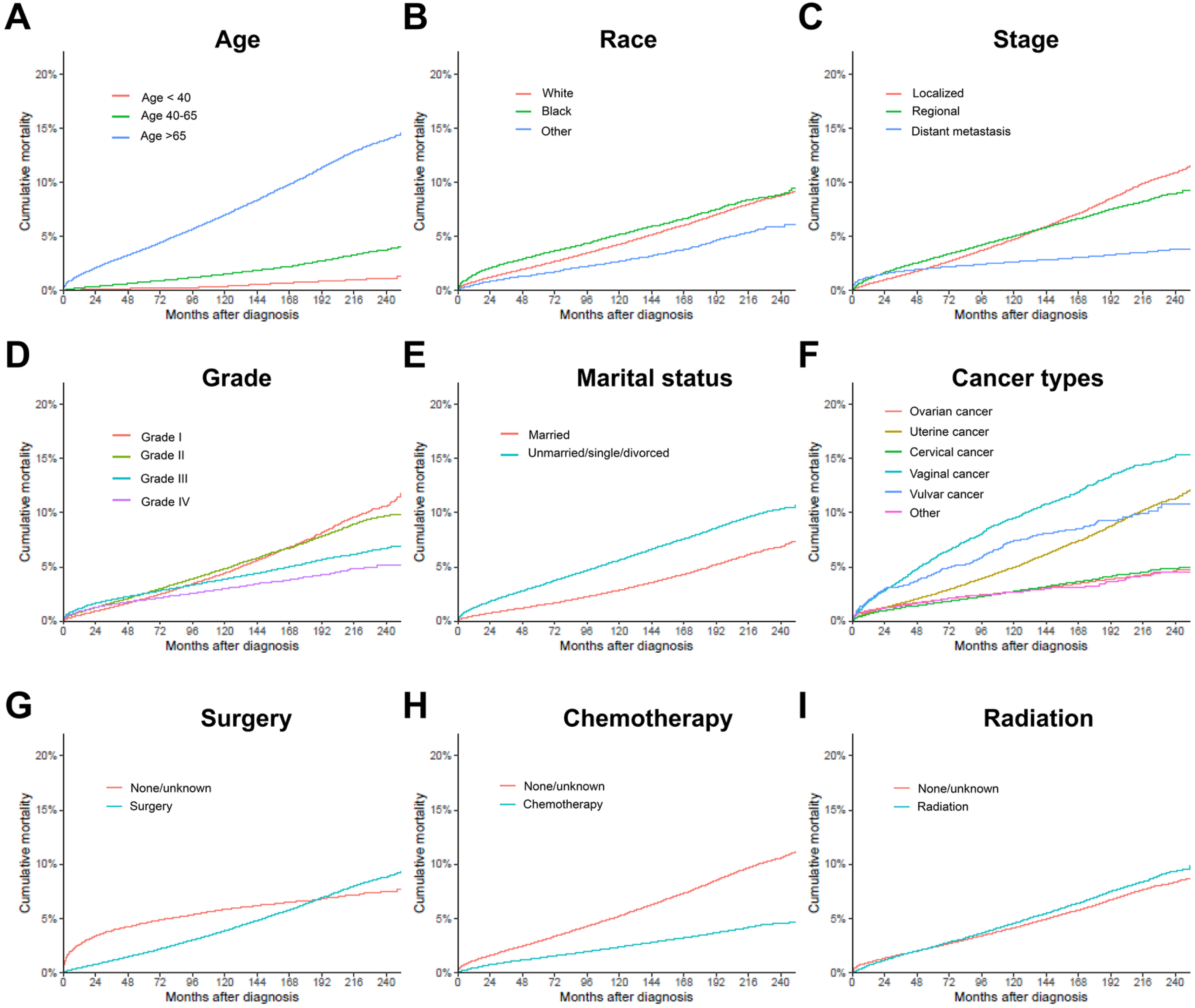
Cumulative Mortality for CVD in Patients with Gynecological Cancers. Cumulative mortality for CVD among gynecological cancer survivors, by age at diagnosis (A), race (B), stage (C), grade (D), marital status at diagnosis (E), cancer types (F), and initial treatment regimens, such as surgery (G), chemotherapy (H), and radiation (I). *Abbreviations: CVD: cardiovascular disease*.

**Table 3.** Univariate and Multivariate Analysis for CVD Death in Patients with Gynecological Cancers. Abbreviations: CVD: cardiovascular disease; HR: hazard ratio; CI: confidence interval. ^a^ malignant neoplasm of other and unspecified female genital organs, or malignant neoplasm of placenta.

| Characteristic | Univariate analysis |  |  | Multivariate analysis |  |  |
| --- | --- | --- | --- | --- | --- | --- |
|  | HR | (95% CI) | *P* value | HR | (95% CI) | *P* value |
| **Age at diagnosis** |  |  | <0.001 |  |  |  |
| <40 |  | ref |  |  | ref |  |
| 40-65 | 3.46 | (2.96, 4.05) |  | 3.71 | (3.17, 4.36) | <0.001 |
| ≥65 | 15.2 | (13.0, 17.7) |  | 15.3 | (13.1, 17.9) | <0.001 |
| **Race** |  |  | <0.001 |  |  |  |
| White |  | ref |  |  | ref |  |
| Black | 1.16 | (1.11, 1.22) |  | 1.12 | (1.07, 1.18) | <0.001 |
| Other | 0.62 | (0.58, 0.67) |  | 0.81 | (0.75, 0.86) | <0.001 |
| **Stage** |  |  | <0.001 |  |  |  |
| Localized |  | ref |  |  | ref |  |
| Regional | 0.98 | (0.94, 1.02) |  | 0.97 | (0.93, 1.01) | 0.200 |
| Distant metastasis | 0.52 | (0.50, 0.54) |  | 0.48 | (0.45, 0.51) | <0.001 |
| **Grade** |  |  | <0.001 |  |  |  |
| Grade I |  | ref |  |  | ref |  |
| Grade II | 1.05 | (1.01, 1.09) |  | 1.1 | (1.06, 1.15) | <0.001 |
| Grade III | 0.83 | (0.80, 0.86) | <0.001 | 1.01 | (0.96, 1.06) | 0.700 |
| Grade IV | 0.59 | (0.55, 0.63) |  | 0.79 | (0.73, 0.84) | <0.001 |
| **Marital status** |  |  |  |  |  |  |
| Married |  | ref |  |  | ref |  |
| Unmarried/Single/Divorced | 1.82 | (1.76, 1.88) |  | 1.57 | (1.52, 1.62) | <0.001 |
| **Cancer type** |  |  | <0.001 |  |  |  |
| Ovarian cancer |  | ref |  |  | ref |  |
| Uterine cancer | 1.9 | (1.82, 1.98) |  | 1.09 | (1.03, 1.15) | 0.003 |
| Cervical cancer | 0.98 | (0.92, 1.04) |  | 0.93 | (0.87, 1.00) | 0.046 |
| Vaginal cancer | 3.39 | (3.18, 3.62) |  | 1.56 | (1.45, 1.68) | <0.001 |
| Vulvar cancer | 2.48 | (2.19, 2.81) |  | 1.12 | (0.98, 1.28) | 0.087 |
| Other<sup>a</sup> | 0.94 | (0.82, 1.08) |  | 0.9 | (0.78, 1.04) | 0.200 |
| **Surgery** |  |  | <0.001 |  |  |  |
| None/unknown |  | ref |  |  | ref |  |
| Yes | 0.72 | (0.69, 0.74) |  | 0.64 | (0.61, 0.67) | <0.001 |
| **Chemotherapy** |  |  | <0.001 |  |  |  |
| None/unknown |  | ref |  |  | ref |  |
| Yes | 0.41 | (0.40, 0.43) |  | 0.59 | (0.56, 0.62) | <0.001 |
| **Radiation** |  |  | <0.001 |  |  |  |
| None/unknown |  | ref |  |  | ref |  |
| Yes | 1.07 | (1.03, 1.11) |  | 0.98 | (0.94, 1.02) | 0.400 |

### Nomogram for CVD Mortality

Patients were randomly divided into the discovery cohort (N = 279,580) and the validation cohort (N = 119,819). The clinical characteristics of patients in the discovery and validation cohorts were summarized in **Table S3**. The nomogram was developed based on the final multivariate model and included the following variables: age at diagnosis, race, stage, grade, marital status, cancer type, and initial treatment regimens (**Figure** 5). The nomogram showed the value for each risk factor, with the total value representing the sum of all variable values. The risk of CVD death could be determined by drawing a line from the total score to the risk score.

**Figure 5.**
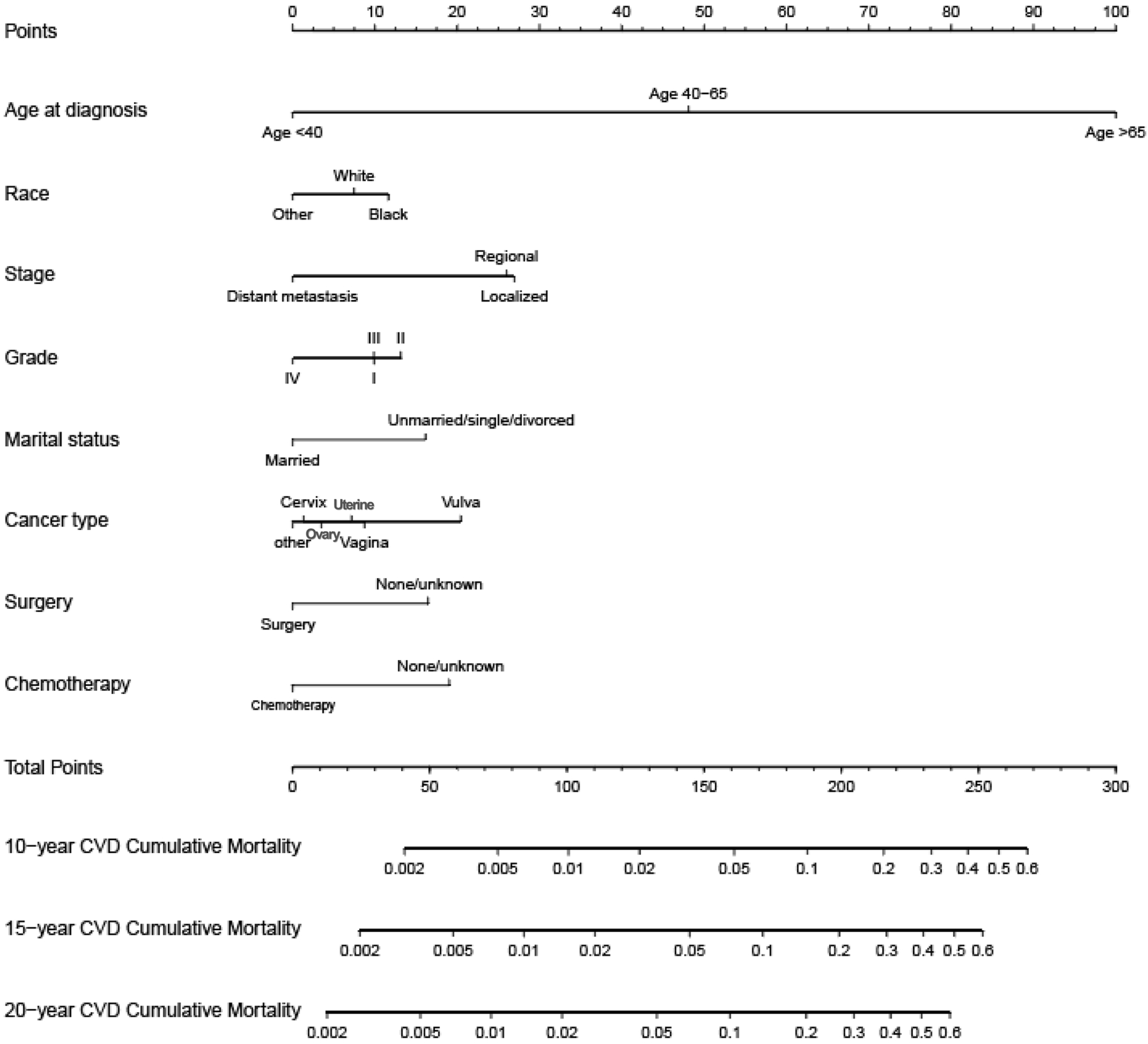
The Nomogram Prediction Model for CVD in Patients with Gynecological Cancers. This nomogram could predict the risk of cardiovascular disease over 10, 15, and 20 years in gynecological cancer patients, using the Fine and Gray competing risk model. Abbreviations: CVD: cardiovascular disease.

In the discovery cohort, the nomogram achieved a C-index of 0.808 (95% CI: 0.784-0.832) for CVD mortality. After validation, the nomogram gained a C-index of 0.797 (95% CI: 0.758-0.837). As shown in the calibration curves, the nomogram achieved a significant agreement between the predicted and actual observations in both the discovery and validation cohorts, as the prediction curves closely aligned with the diagonal. The AUC values in the discovery cohort for 20-year CVD mortality were 74.9% (**Figure S2**). After validation, the nomogram achieved an AUC of 75.7% for 20-year CVD death (**Figure S2**). These results suggested a satisfactory potential clinical effect of this model for predicting CVD mortality risk in patients with GCs. To test the agreement between the real and ideal values of the model, the calibration plot was utilized to confirm the discrimination ability. The calibration plots for 10-, 15-, and 20-year CVD death probabilities closely aligned with the standard line, indicating a high level of calibration (**Figure S3**). A DCA curve was subsequently utilized to illustrate the clinical validity of the nomogram (**Figure S4**).

## DISCUSSION

In recent years, survival rates for GCs have markedly improved ^42–44^. This study investigated CVD mortality risk in GC patients and found it higher than the general US population, particularly in younger patients. Factors such as age at diagnosis, ethnicity, cancer types, and treatment modalities were associated with the increased CVD death risk. A predictive model was developed to assess the CVD mortality risk in GC patients, emphasizing the importance of cardiovascular health in improving long-term survival rates.

Women encountered an elevated risk of both cardiovascular disease and cancer, especially as they aged ^21, 45–48^. Both CVD and specific types of cancer had common risk factors including smoking, physical inactivity, obesity, diabetes, hypertension, and hypercholesterolemia^21, 49^. Physical inactivity was associated with increased risks of both CVD and cancer, whereas highly active women exhibited a lower risk of endometrial cancer^50–54^. Notably, women with endometrial cancer exhibited an increased risk of CVD^55^. Obesity was correlated with elevated risks of endometrial and breast cancers, as well as increased cancer-related morbidity and mortality^56–59^. This research found that patients with GCs had a CVD mortality rate of 10.0%. Remarkably, younger cancer survivors, particularly those diagnosed before age 45, had a higher risk of mortality from CVD compared to the general US population. Among GC patients, older age and being unmarried, single, or divorced were linked to higher CVD death risk, whereas treatments involving surgery or chemotherapy were associated with lower risk. These findings underscored the importance of addressing common risk factors and delivering comprehensive care for women to confront both cancer and cardiovascular diseases.

GCs accounted for 11% of all cancers in women, encompassing endometrial, ovarian, and cervical cancers^1^. Advances in detection and treatment had significantly improved outcomes for advanced and recurrent GC cases^4–8, 42, 60^. However, treatments were associated with risks such as therapy-related cardiovascular toxicity, a growing concern in oncology^61, 62^. The interplay between cardiovascular care and GC survivors was increasingly critical due to their elevated risk of CVD mortality from cancer treatments^9, 11, 15, 49, 55, 63–66^. Various treatments for GCs carried distinct CVD risks^42, 67–71^. Anthracyclines, including doxorubicin and its pegylated liposomal form, were crucial for treating advanced ovarian and endometrial cancers but exhibited dose-dependent cardiotoxicity^72–74^. Pegylated liposomal doxorubicin (PLD) had lower cardiotoxicity compared to doxorubicin and was utilized in the treatment of gynecologic malignancies and uterine sarcomas^75^. Platinum-based drugs, such as cisplatin, commonly used for cervical, endometrial, and ovarian cancers, increased the risk of thromboembolic events and cardiovascular complications^76–79^. Paclitaxel and docetaxel, frequently employed in the treatment of cervical, endometrial, and ovarian cancers, might induce arrhythmias^80–84^. Antimetabolites, such as fluorouracil (5-FU), commonly used for cervical and ovarian cancers, could precipitate heart problems, particularly in patients with pre-existing coronary artery disease^85–87^. Antiangiogenic drugs, including bevacizumab and pazopanib, used to treat cervical, epithelial, and mucinous ovarian cancers, as well as leiomyosarcoma, were linked to cardiovascular issues such as hypertension, heart failure, and arterial thromboembolic events^88–91^. Immune checkpoint inhibitors, approved for the treatment of uterine and cervical cancers, could cause rare but serious cardiac complications, including myocarditis and acute myocardial infarction ^92–94^. Tamoxifen, frequently used in the treatment of endometrial cancer, could exert estrogen-like effects on the heart, potentially leading to fatal long-term complications related to serious blood clots^95, 96^. Additionally, women with GCs might experience unique consequences of cancer treatment, such as premature menopause and reduced fertility^97–99^. Factors such as ovarian reserve, exposure to cancer treatments, and duration of therapy could influence the onset of early menopause ^100, 101^. These factors elevated the risk of coronary artery disease, worsened cancer patient outcomes, and heightened the likelihood of cardiovascular diseases, heart failure, and stroke, primarily due to the prolonged absence of estrogen affecting metabolic and vascular function^101–103^.

Various strategies have been investigated to mitigate the risk of cardiotoxicity in patients undergoing treatment for GCs, emphasizing the customization of approaches based on individual cardiovascular risk factors^104^. These strategies included avoiding cardiotoxic treatments when feasible, considering alternative therapies, and consulting with a multidisciplinary team before initiating treatments ^104, 105^. For instance, in the case of doxorubicin treatment, potential strategies included adjusting infusion schedules, using dexrazoxane, or opting for liposomal formulations^106,107^. In ovarian cancer, pegylated liposomal doxorubicin (PLD) was commonly employed in cardioprotective regimens ^108, 109^. Additionally, dose reduction or modification of the infusion schedule could help alleviate cardiotoxicity associated with 5-fluorouracil (5-FU)^110^. The nomogram had been developed to predict the CVD mortality risk in patients with GCs, providing valuable guidance.

The study encountered several limitations. The primary concern was the potential introduction of bias due to the retrospective nature of the data derived from the SEER database. Inadequate information on cardiovascular complications, risk factors, disease history, and therapeutic methods hindered the assessment of their impact on CVD death risk. Additionally, the absence of crucial factors such as genetic markers, behavioral patterns, and other characteristics limited the study’s accuracy. To address these limitations, future cohort studies should prioritize identifying prognostic markers and incorporating a wider range of factors to enhance precision.

## CONCLUSIONS

This research underscored the heightened risk of CVD mortality faced by individuals with GCs, particularly among younger survivors. Various factors including age, ethnicity, cancer type, and treatment approaches contributed to this risk. As a result, a predictive model was created to evaluate the likelihood of CVD mortality in patients with GCs, emphasizing the significance of prioritizing heart health to improve survival. These findings underscored the necessity for personalized interventions and proactive management of cardiovascular risk factors in individuals with GCs, with the ultimate goal of reducing the impact of cardiovascular disease.

## Acknowledgments

The data used in this study was from the Surveillance, Epidemiology, and End Results (SEER) Registry database.

## Contributors

YY: conceptualization, methodology, software, formal analysis, writing – original draft, writing – review and editing, guarantor. YJP: conceptualization, methodology, writing – original draft, writing – review and editing, guarantor. LBS: methodology, formal analysis, writing – review and editing, supervision. CLW: conceptualization, methodology, writing – review and editing, supervision. WSJ: conceptualization, methodology, project administration, writing – review and editing, supervision. CYG: conceptualization, methodology, data curation, project administration, funding acquisition, writing – review and editing, supervision.

## Patient consent for publication

Not applicable.

## Provenance and peer review

Not commissioned; externally peer reviewed.

## Data availability statement

Data may be obtained from a third party and are not publicly available.

## Supplemental material

This content has been supplied by the author(s). It has not been vetted by BMJ Publishing Group Limited (BMJ) and may not have been peer-reviewed. Any opinions or recommendations discussed are solely those of the author(s) and are not endorsed by BMJ. BMJ disclaims all liability and responsibility arising from any reliance placed on the content. Where the content includes any translated material, BMJ does not warrant the accuracy and reliability of the translations (including but not limited to local regulations, clinical guidelines, terminology, drug names, and drug dosages), and is not responsible for any error and/or omissions arising from translation and adaptation or otherwise.

