## supplementary material for "Cardiovascular Mortality in Patients with Gynecological Cancers: A Population-based Cohort Study"

**Table S1**. **Cumulative Incidence of CVD Mortality.**

| **Characteristic** | **10-year CVD**  **death rate** | **15-year CVD**  **death rate** | **20-year CVD**  **death rate** |
| --- | --- | --- | --- |
| **Age at diagnosis** |  |  |  |
| <40 | 0.37% (0.30%, 0.46%) | 0.78% (0.65%, 0.92%) | 1.1% (0.93%, 1.4%) |
| 40-65 | 1.5% (1.4%, 1.6%) | 2.5% (2.4%, 2.6%) | 3.8% (3.6%, 4.0%) |
| ≥65 | 7.0% (6.8%, 7.1%) | 11% (10%, 11%) | 14% (14%, 14%) |
| **Race** |  |  |  |
| White | 4.3% (4.2%, 4.3%) | 6.5% (6.4%, 6.6%) | 8.8% (8.6%, 8.9%) |
| Black | 5.2% (4.9%, 5.4%) | 7.1% (6.7%, 7.4%) | 8.9% (8.4%, 9.5%) |
| Other | 2.7% (2.5%, 2.9%) | 4.2% (3.9%, 4.5%) | 5.9% (5.4%, 6.4%) |
| **Stage** |  |  |  |
| Localized | 4.7% (4.6%, 4.8%) | 7.8% (7.6%, 8.0%) | 11% (11%, 11%) |
| Regional | 5.0% (4.8%, 5.2%) | 7.1% (6.8%, 7.3%) | 9.0% (8.6%, 9.3%) |
| Distant metastasis | 2.6% (2.5%, 2.7%) | 3.2% (3.0%, 3.3%) | 3.8% (3.6%, 4.0%) |
| **Grade** |  |  |  |
| Grade I | 4.4% (4.3%, 4.6%) | 7.5% (7.2%, 7.7%) | 11% (10%, 11%) |
| Grade II | 4.8% (4.7%, 5.0%) | 7.3% (7.1%, 7.5%) | 9.7% (9.4%, 10%) |
| Grade III | 3.9% (3.7%, 4.0%) | 5.3% (5.2%, 5.5%) | 6.7% (6.5%, 7.0%) |
| Grade IV | 3.0% (2.8%, 3.2%) | 4.1% (3.8%, 4.4%) | 5.2% (4.7%, 5.7%) |
| **Marital status** |  |  |  |
| Married | 2.9% (2.8%, 2.9%) | 4.8% (4.6%, 4.9%) | 6.9% (6.6%, 7.1%) |
| Unmarried/Single/Divorced | 5.6% (5.5%, 5.7%) | 8.1% (7.9%, 8.2%) | 10% (10%, 11%) |
| **Cancer type** |  |  |  |
| Ovarian cancer | 2.7% (2.6%, 2.9%) | 3.7% (3.5%, 3.8%) | 4.6% (4.4%, 4.9%) |
| Uterine cancer | 5.0% (4.8%, 5.1%) | 8.1% (7.9%, 8.2%) | 11% (11%, 12%) |
| Cervical cancer | 2.7% (2.6%, 2.9%) | 3.9% (3.7%, 4.1%) | 4.9% (4.6%, 5.2%) |
| Vaginal cancer | 9.5% (9.0%, 10%) | 13% (12%, 13%) | 15% (14%, 16%) |
| Vulvar cancer | 7.4% (6.5%, 8.4%) | 9.0% (7.9%, 10%) | 11% (9.3%, 12%) |
| Other | 2.7% (2.3%, 3.1%) | 3.3% (2.8%, 3.8%) | 4.5% (3.7%, 5.4%) |
| **Surgery** |  |  |  |
| None/unknown | 5.8% (5.6%, 6.0%) | 6.6% (6.4%, 6.8%) | 7.5% (7.2%, 7.8%) |
| Yes | 3.8% (3.8%, 3.9%) | 6.3% (6.2%, 6.5%) | 8.8% (8.6%, 9.0%) |
| **Chemotherapy** |  |  |  |
| None/unknown | 5.2% (5.1%, 5.3%) | 7.9% (7.8%, 8.0%) | 11% (10%, 11%) |
| Yes | 2.4% (2.3%, 2.5%) | 3.4% (3.3%, 3.6%) | 4.5% (4.3%, 4.8%) |
| **Radiation** |  |  |  |
| None/unknown | 4.1% (4.0%, 4.2%) | 6.2% (6.1%, 6.3%) | 8.3% (8.1%, 8.5%) |
| Yes | 4.5% (4.4%, 4.7%) | 6.9% (6.7%, 7.2%) | 9.3% (8.9%, 9.7%) |

Abbreviations: CVD: cardiovascular disease.

**Table S2. Average Annual Percentage Change of CVD Death in Patients with Gynecological Cancers.**

| **Cancer types** | **2000 Year SMR (95% CI)** | **2020 Year SMR (95% CI)** | **2000-2020 Year AAPC** |
| --- | --- | --- | --- |
| Gynecological cancers | 1.109 (1.055-1.166) | 1.655 (1.207-2.214) | 0.0262 (0.0220-0.0304) * |
| Ovarian cancer | 0.963 (0.836-1.103) | 2.249 (1.123-4.024) | 0.0668 (0.0537-0.0798) * |
| Uterine cancer | 1.063 (0.998-1.131) | 1.542 (0.988-2.294) | 0.0229 (0.0184-0.0274) * |
| Cervical cancer | 1.482 (1.277-1.711) | 1.919 (0.523-4.913) | 0.0417 (0.027-0.0564) * |
| Vaginal cancer | 1.878 (1.248-2.715) | 3.954 (0.479-14.282) | 0.1071 (0.0662-0.1479) * |
| Vulvar cancer | 1.354 (1.114-1.629) | 0.858 (0.104-3.098) | -0.0239 (-0.0413--0.0064) |

AAPC was a summary measure that used a single number to describe the average APCs over multiple years, even when the changes in trends were indicated by the joinpoint model. It was computed as a weighted average of the APCs from the joinpoint model, with weights equal to the length of the APC interval. *Abbreviations: CVD: cardiovascular disease; AAPC: average annual percentage change; CI: confidence interval; ^a*^ indicated that 95% confidence interval does not cross 0 and the AAPC was significant.*

**Table S3**. **Participant Characteristics in the Discovery and Validation Cohorts.**

| **Characteristic** | **Discovery cohort**  **(N = 279,580)** | **Validation cohort**  **(N = 119,819)** |
| --- | --- | --- |
| **Age at diagnosis** |  |  |
| <40 | 24,832 (8.9%) | 10,642 (8.9%) |
| 40-65 | 106,702 (38%) | 45,729 (38%) |
| ≥65 | 148,046 (53%) | 63,448 (53%) |
| **Race** |  |  |
| White | 225,168 (81%) | 96,742 (81%) |
| Black | 28,152 (10%) | 11,813 (9.9%) |
| Other | 26,260 (9.4%) | 11,264 (9.4%) |
| **Stage** |  |  |
| Localized | 146,762 (52%) | 63,036 (53%) |
| Regional | 58,067 (21%) | 24,633 (21%) |
| Distant metastasis | 74,751 (27%) | 32,150 (27%) |
| **Grade** |  |  |
| Grade I | 81,857 (29%) | 35,093 (29%) |
| Grade II | 79,208 (28%) | 33,485 (28%) |
| Grade III | 85,937 (31%) | 37,084 (31%) |
| Grade IV | 32,578 (12%) | 14,157 (12%) |
| **Marital status** |  |  |
| Married | 140,697 (50%) | 60,377 (50%) |
| Unmarried/Single/Divorced | 138,883 (50%) | 59,442 (50%) |
| **Cancer type** |  |  |
| Ovarian cancer | 66,061 (24%) | 28,502 (24%) |
| Uterine cancer | 147,556 (53%) | 63,108 (53%) |
| Cervical cancer | 44,839 (16%) | 19,186 (16%) |
| Vaginal cancer | 11,509 (4.1%) | 4,861 (4.1%) |
| Vulvar cancer | 2,898 (1.0%) | 1,278 (1.1%) |
| Other | 6,717 (2.4%) | 2,884 (2.4%) |
| **Surgery** |  |  |
| None/unknown | 55,430 (20%) | 23,746 (20%) |
| Yes | 224,150 (80%) | 96,073 (80%) |
| **Chemotherapy** |  |  |
| None/unknown | 180,382 (65%) | 77,296 (65%) |
| Yes | 99,198 (35%) | 42,523 (35%) |
| **Radiation** |  |  |
| None/unknown | 211,161 (76%) | 90,767 (76%) |
| Yes | 68,419 (24%) | 29,052 (24%) |
| **Cause of death** |  |  |
| Alive | 165,364 (59%) | 70,894 (59%) |
| CVD | 11,324 (4.1%) | 5,047 (4.2%) |
| Cancer-index | 82,236 (29%) | 35,315 (29%) |
| Cancer-non-index | 4,910 (1.8%) | 2,074 (1.7%) |
| No-CVD other cause | 15,746 (5.6%) | 6,489 (5.4%) |

*Abbreviations: CVD: cardiovascular disease.*

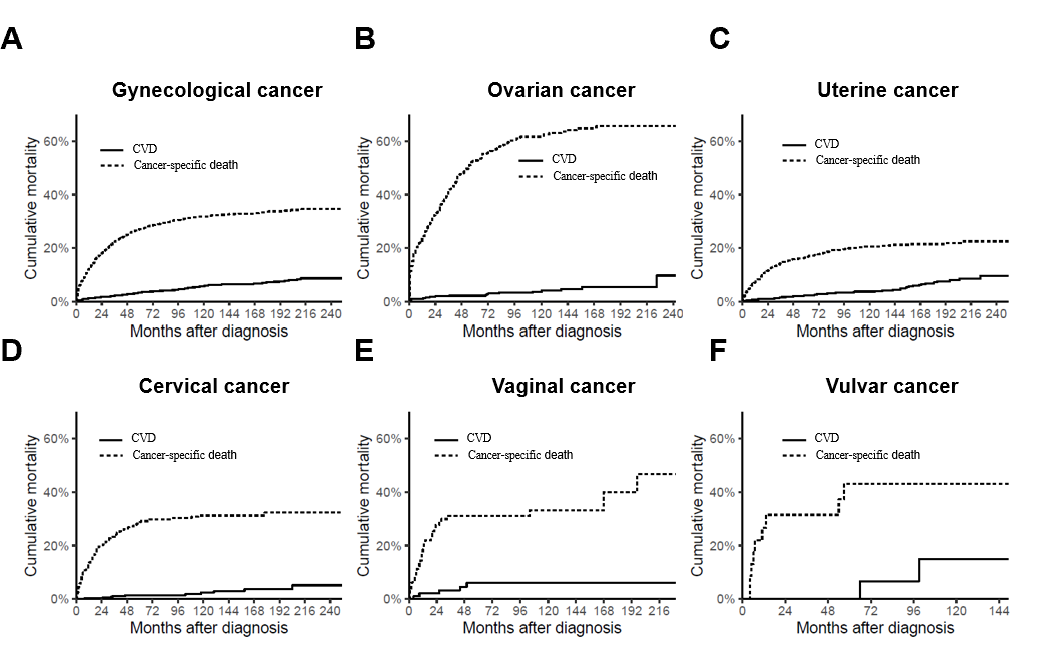

**Figure S1**. **Cumulative Mortality for Cancer-Specific Death and CVD in Patients with Gynecological Cancers.**

*Abbreviations: CVD: cardiovascular disease.*

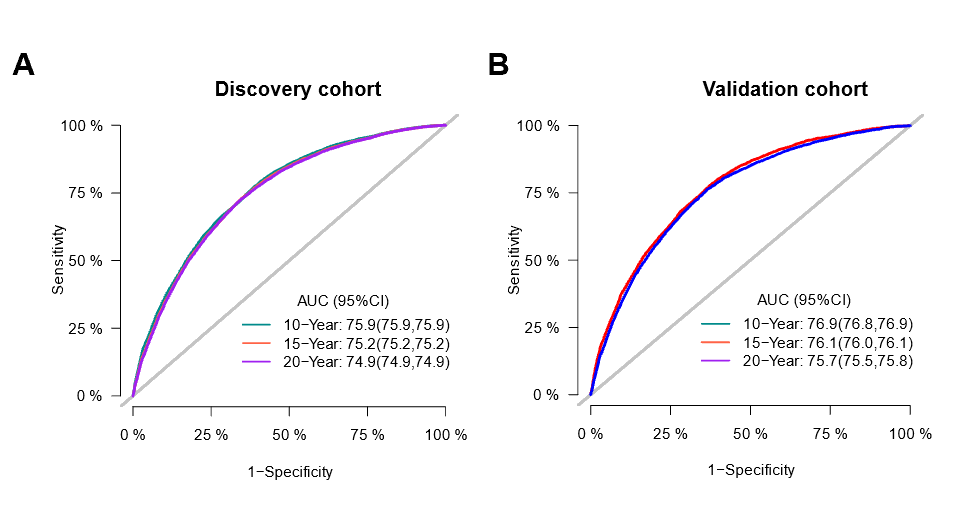

**Figure S2**. **ROC Curves for the Nomogram of CVD in the Discovery (A) and Validation Cohort (B).**

*Abbreviations: CVD: cardiovascular disease; ROC: receiver operator characteristic.*

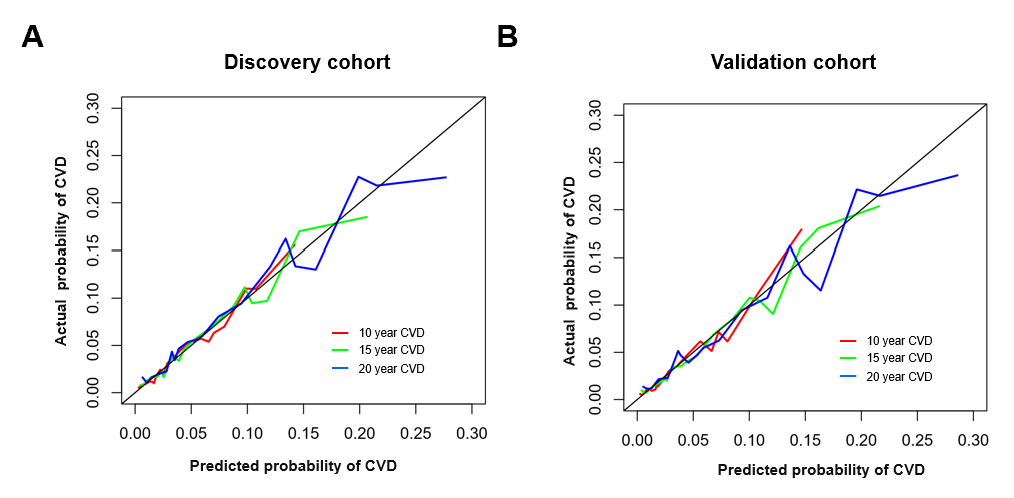

**Figure S3**. **Calibration plots for the Nomogram of CVD in the Discovery (A) and Validation Cohort (B).**

The perfect predictions were shown by the 45° line, demonstrating a high level of similarity between the predicted and actual CVD death rates. *Abbreviations: CVD: cardiovascular disease.*

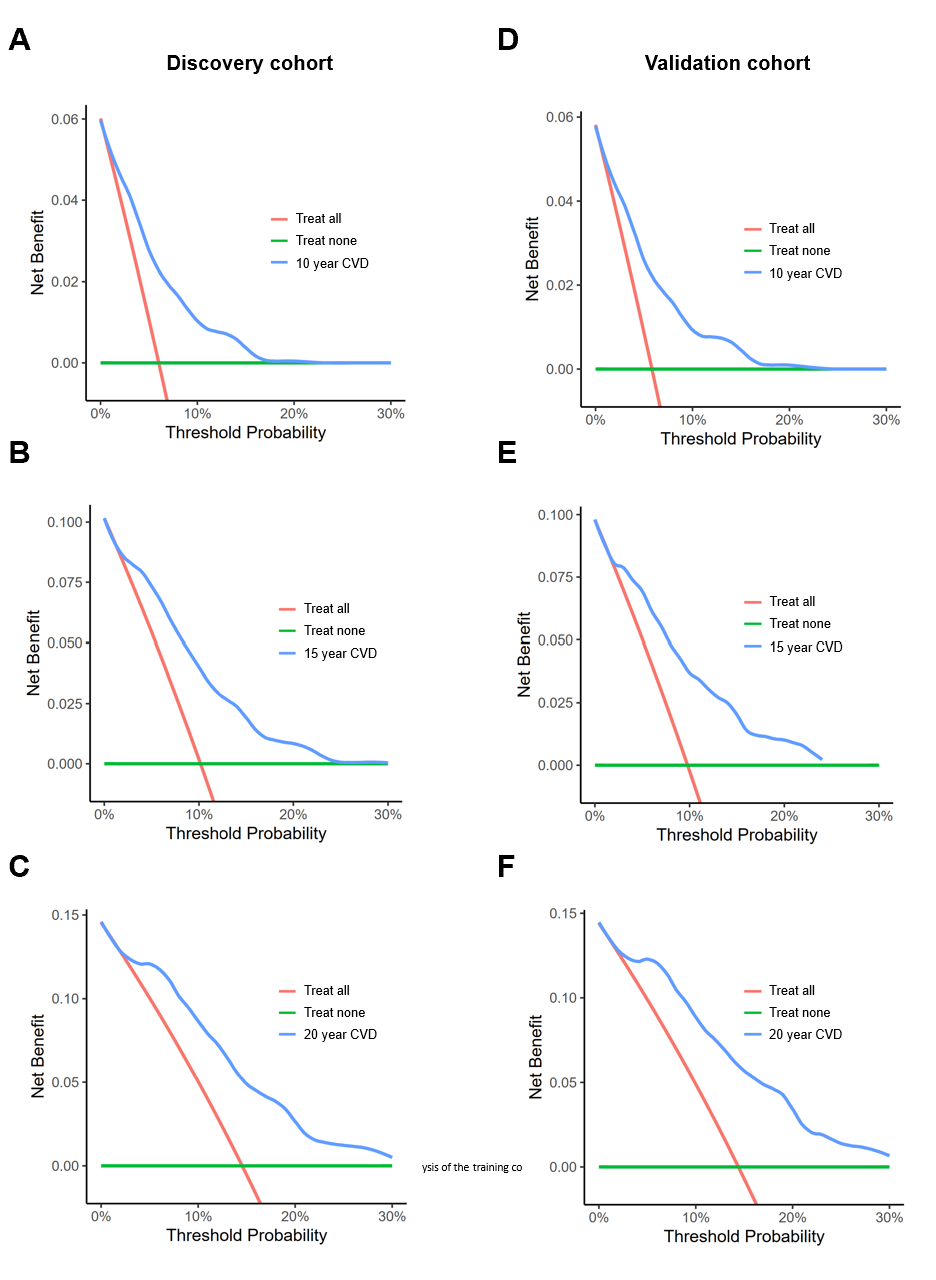

**Figure S4. Decision Curve Analyses for the Nomogram of CVD in the Discovery (A-C) and Validation Cohort (D-F).**

The green line suggested that patients should not take the necessary measures, while the red line demonstrated that all patients should. The y-axis represented the net benefit by adding benefit points and subtracting harm points. Our findings indicated the net benefit offered by the nomogram (blue line) in the discovery (A-C) and validation (D-F) cohorts. Abbreviations: CVD: cardiovascular disease.
